# Early life infection and proinflammatory, atherogenic metabolomic and lipidomic profiles at 12 months of age: a population-based cohort study

**DOI:** 10.1101/2021.12.02.21267173

**Authors:** Toby Mansell, Richard Saffery, Satvika Burugupalli, Anne-Louise Ponsonby, Mimi LK Tang, Martin O’Hely, Siroon Bekkering, Adam AT Smith, Rebecca Rowland, Sarath Ranganathan, Peter D Sly, Peter Vuillermin, Fiona Collier, Peter J Meikle, David P Burgner, on behalf of the Barwon Infant Study Investigator Group

## Abstract

**Background:** The risk of adult onset cardiovascular and metabolic (cardiometabolic) disease accrues from early life. Infection is ubiquitous in infancy and induces inflammation, a key cardiometabolic risk factor, but the relationship between infection, inflammation, and metabolic profiles in early childhood remains unexplored. We investigated relationships between infection and plasma metabolomic and lipidomic profiles at age 12 months, and mediation of these associations by inflammation.

**Methods:** Matched infection, metabolomics and lipidomics data were generated from 555 infants in a pre-birth longitudinal cohort. Infection data from birth to 12 months were parent-reported (total infections at age 1, 3, 6, 9, and 12 months), inflammation markers (high-sensitivity C-reactive protein, hsCRP); glycoprotein acetyls GlycA) were quantified at 12 months. Metabolic profiles were 12-month plasma nuclear magnetic resonance metabolomics (228 metabolites) and liquid-chromatography/mass-spectrometry lipidomics (776 lipids). Associations were evaluated with multivariable linear regression models.

**Results:** Frequent infant infections were associated with adverse metabolomic (elevated inflammation markers, triglycerides, phenylalanine, and lower HDL cholesterol, apolipoprotein A1, and omega-3 fatty acids) and lipidomic profiles (elevated phosphatidylethanolamines and lower hexosylceramides, trihexosylceramides, and cholesteryl esters). Similar, more marked, profiles were observed with higher GlycA, but not hsCRP. GlycA, but not hsCRP, mediated a substantial proportion of the relationship between infection and metabolome/lipidome.

**Conclusions:** Infants with a greater infection burden from birth to 12 months had pro-inflammatory and pro-atherogenic plasma metabolomic/lipid profiles, indicative of heightened risk of cardiovascular disease, obesity, and type 2 diabetes in adults. These findings suggest potentially modifiable pathways linking early life infection and inflammation with subsequent cardiometabolic risk.

**Funding:** The establishment work and infrastructure for the BIS was provided by the Murdoch Children’s Research Institute (MCRI), Deakin University and Barwon Health. Subsequent funding was secured from National Health and Medical Research Council of Australia (NHMRC), The Shepherd Foundation, The Jack Brockhoff Foundation, the Scobie & Claire McKinnon Trust, the Shane O’Brien Memorial Asthma Foundation, the Our Women’s Our Children’s Fund Raising Committee Barwon Health, the Rotary Club of Geelong, the Minderoo Foundation, the Ilhan Food Allergy Foundation, GMHBA, Vanguard Investments Australia Ltd, and the Percy Baxter Charitable Trust, Perpetual Trustees. In-kind support was provided by the Cotton on Foundation and CreativeForce. The study sponsors were not involved in the collection, analysis, and interpretation of data; writing of the report; or the decision to submit the report for publication. Research at MCRI is supported by the Victorian Government’s Operational Infrastructure Support Program. This work was also supported by NHMRC Senior Research Fellowships (1008396 to ALP; 1064629 to DB; 1045161 to RS), NHMRC Investigator Grants to ALP (1110200) and DB (1175744), NHMRC-A*STAR project grant (1149047). TM is supported by an MCRI ECR Fellowship. SB is supported by the Dutch Research Council (452173113).

## Introduction

Infectious diseases are ubiquitous in infancy and childhood, with potential long-term impacts on health across the life course. Infection has been recognised as a potential contributor to atherosclerotic cardiovascular disease (CVD), one of the leading causes of adult morbidity and mortality, since the 19^th^ century(Nieto, 1998). More recent adult studies link previous infection with long-term risks of disease(Bergh et al., 2017; Cowan et al., 2018; H. Wang et al., 2017). The mechanisms are largely unknown, but likely include immune activation and heightened inflammation (Shah, 2019), which are pathways central to CVD pathogenesis(Donath, Meier, & Böni-Schnetzler, 2019; Ferrucci & Fabbri, 2018) and therefore offer potentially druggable targets in high-risk individuals(Donath, Dinarello, & Mandrup-Poulsen, 2019; Ridker et al., 2017). Glycoprotein acetyls (GlycA), an NMR composite measure of cumulative, chronic inflammation(Connelly, Otvos, Shalaurova, Playford, & Mehta, 2017), is an emerging biomarker for cardiometabolic risk(Connelly et al., 2017) that out-performs high-sensitivity C-reactive protein (hsCRP) as a predictor of CVD events and mortality(Akinkuolie, Buring, Ridker, & Mora, 2014; Duprez et al., 2016), and of infection-related morbidity and mortality(Ritchie et al., 2015).

Cardiovascular and metabolic (cardiometabolic) disease pathogenesis begins in early life and accrues across the life course(Nakashima, Wight, & Sueishi, 2008). Infections occur disproportionally in early childhood(Cromer et al., 2014; Grüber et al., 2008; Troeger et al., 2018; Tsagarakis et al., 2018), and there is a dose-response relationship between childhood infections, adverse cardiometabolic phenotypes(D. P. Burgner, M. A. Sabin, et al., 2015) and CVD events(D. P. Burgner, M. N. Cooper, et al., 2015) in adulthood. Infection is linked to pro-atherogenic metabolic perturbations in later childhood and adulthood(Feingold & Grunfeld, 2019; Khovidhunkit, Memon, Feingold, & Grunfeld, 2000), including higher triglycerides, lower high-density lipoprotein (HDL) cholesterol and apolipoprotein A1 (ApoA1), and oxidised low-density lipoprotein (LDL)(Liuba, Persson, Luoma, Ylä-Herttuala, & Pesonen, 2003; Pesonen, Rapola, Viikari, Turtinen, & Akerblom, 1993), and to acute and chronic inflammation(Burgner, Liu, Wake, & Uiterwaal, 2015; Ritchie et al., 2015), but little is known about these relationships in early life, when most infections occur.

We therefore aimed to characterise metabolomic and lipidomic profiles at 12 months of age and their relationship to infection burden in the first year of life. We also investigated the extent to which inflammation mediated the relationship between infection burden and metabolomic and lipidomic differences.

## Methods

### Study cohort

This study used available data from 555 mother-infant dyads in the Barwon Infant Study (BIS), a population-based pre-birth longitudinal cohort (n=1,074 mother-infant dyads). The cohort details and inclusion/exclusion criteria have been detailed elsewhere(Vuillermin et al., 2015); in brief mothers were eligible if they were residents of the Barwon region in south-east Australia and planned to give birth at the local public or private hospital. Mothers were recruited at approximately 15 weeks’ gestation and provided informed consent. They were excluded if they were not a permanent Australian resident, aged <18 years, required an interpreter to complete questionnaires, or had previously participated in BIS. Infants were excluded if they were very preterm (<32 completed weeks gestation) or had a serious illness or major congenital malformation identified during the first few days of life. Ethics approval was granted by the Barwon Health Human Research Ethics Committee (HREC 10/24).

### Parent-reported infections

At the 4-week, 3-month, 6-month, 9-month and 12-month time points following birth, mothers were asked to report each episode of infant illness or infection since the previous timepoint using standardised on-line questionnaires. The number of parent-reported infections from birth to 12 months was defined as the total number of respiratory tract infections, gastroenteritis, conjunctivitis and acute otitis media episodes from birth to the 12-month timepoint. It was not possible to identify the proportion of parent-reported infections that lead to health service utilisation(Rowland et al., 2020).

### Other maternal and infant measures

Questionnaires during pregnancy and at birth were used to collect self-reported data on maternal age, household income, and prenatal smoking (considered here as a dichotomous any/none exposure). Infant gestational age and birth weight were collected from birth records, and the age- and sex-standardised birth weight z-score was calculated using the 2009 revised British United Kingdom World Health Organisation (UK-WHO) growth charts(Cole, Williams, & Wright, 2011).

Breastfeeding duration up to 12 months of age was collected from maternal questionnaire data. As most evidence for the protective effect of breastfeeding on early life infection is from comparisons between any breastfeeding and no breastfeeding(Victora et al., 2016), and in light of previous evidence in BIS for an association between even a short duration of breastfeeding and lower odds of infection in early infancy(Rowland et al., 2020), we first looked at breastfeeding as a binary (any/none) measure in models (presented in the main text). As most infants (98.2%) were breastfed to some extent, and it is unknown the degree to which breastfeeding, and the timing of breastfeeding, might affect 12-month metabolomics and lipidomics, we also considered duration of breastfeeding to 12 months of age for sensitivity analyses (see Supplementary Files 1C, 1D, 1E, 1G, 1H, 1I). Data on mode of birth was collected from birth records and categorised as vaginal, planned, or unplanned caesarean section delivery.

### Metabolomic and lipidomic profiling

Venous peripheral blood was collected from infants at the 12-month time point in sodium heparin and generally processed within 4 hours, with a minority (197 of 555) processed after 4 hours (median time for those 197 samples = 19.9 hours, IQR [18.7, 21.4]). Sample time to post-processing storage was included as a covariate in analyses. As a sensitivity analysis, participants with a plasma storage time greater than 4 hours (197 out of 555 infants) were excluded, and this had little difference on the estimated effect sizes observed in analyses with the full cohort (Supplementary Files 1A and 1B). Plasma was stored at −80°C, and aliquots were shipped on dry ice to Nightingale Health (Helsinki, Finland) for NMR metabolomic quantification and Baker IDI (Melbourne, Australia) for LC/MS lipidomic quantification as described below. For secondary analyses investigating possible ‘reverse causality’, i.e., whether metabolomic or lipidomic profile at birth was associated with number of parent-reported infections from birth to 12 months of age, metabolomics and lipidomics data using the same platforms from venous cord blood collected at birth was used.

The NMR-based metabolomics platform has been described in detail (Kettunen et al., 2016; Soininen, Kangas, Würtz, Suna, & Ala-Korpela, 2015), and quantified a broad range of metabolic measures including lipoprotein size subclasses, triglycerides, cholesterols, fatty acids, amino acids, ketone bodies, glycolysis metabolites, and glycoprotein acetyls (GlycA), a measure of acute and chronic inflammation(Connelly et al., 2017). In brief, plasma samples were mixed with a sodium phosphate buffer prior to NMR measurements with Bruker AVANCE III 500 MHz and Bruker AVANCE III HD 600 MHz spectrometers (Bruker, Billerica, Massachusetts). Samples were kept at 6°C using the SampleJet sample changer (Bruker) to prevent degradation. After initial measurements, samples went through a multiple-step lipid extraction procedure using saturated sodium chloride solution, methanol, dichloromethane, and deuterochloroform. The lipid extracts were then analysed using the 600 MHz instrument(Soininen et al., 2015). The utility of this platform in epidemiological research has been detailed elsewhere(Würtz et al., 2017). Using the Nightingale Health 2016 bioinformatics protocol, 228 metabolomic measures were generated. From a subset of participants, replicate samples were quantified, and these showed a low percentage coefficient of variation (<10%). As a large proportion of these measures are ratios and are strongly correlated with each other in children and adults(Ellul et al., 2019), an informative subset of 51 measures that captured the majority of variation the metabolomic dataset, primarily absolute metabolite concentrations, were included in analysis presented in the main text.

Analyses for excluded metabolomic measures are presented as supplementary data. To complement GlycA as a measure of inflammation, high sensitivity C-reactive protein (hsCRP) was also quantified in 12-month plasma using ELISA assay (R&D Systems, Minneapolis, Minnesota, cat. no. DY1707), as per manufacturer instruction.

The details of the high-performance LC/MS lipidomics platform have been described elsewhere(Beyene et al., 2020). In addition, we used medronic acid to passivate the LC/MS system to avoid peak tailing for acidic phospholipids(Hsiao, Potter, Chu, & Yin, 2018). In brief, this platform quantified 776 lipid features in 36 lipid classes, including sphingolipids, glycerophospholipids, sterols, glycerolipids and fatty acyls. Analysis was performed on an Agilent 6490 QQQ mass spectrometer with an Agilent 1290 series HPLC system and two ZORBAX eclipse plus C18 column (2.1×100mm 1.8mm) (Agilent, Santa Clara, California) with the thermostat set at 45°C. Mass spectrometry analysis was performed in both positive and negative ion mode with dynamic scheduled multiple reaction monitoring (MRM).

Quantification of lipid species was determined by comparison to the relevant internal standard. Lipid class total concentrations were calculated as the sum of individual lipid species concentrations, except in the case of TGs and TG-Os, where we measured both neutral loss [NL] and single ion monitoring [SIM] peaks, and subsequently used the [SIM] species concentrations for summation purposes.

### Statistical analysis

Analyses were performed in R (version 3.6.3)(Team, 2018). All metabolomic and lipidomic measures had 1 added to their value before they were natural log-transformed and scaled to a standard distribution (standard deviation units). Pearson’s correlations were calculated for number of infections from birth to 12 months with 12-month GlycA and hsCRP.

The estimated effect of number of parent-reported infections as an exposure on metabolomic and lipidomic profile was investigated using linear regression models for each metabolomic/lipidomic measure. Standard error was used to calculate 95% confidence intervals for estimated effects. All models were adjusted for infant sex, exact age at 12-month time point, birth weight z-score, maternal household income, exposure to maternal smoking during pregnancy, breastfeeding, and time from collection to storage for 12-month plasma samples. Linear regression models adjusted for the same covariates were used to investigate GlycA and hsCRP as exposures and each metabolomic/lipidomic measure as an outcome. P-values are two-tailed, and were adjusted for multiple comparisons within each dataset (NMR metabolomics, LC/MS lipidomic species, and LC/MS lipidomic classes) using the Benjamini-Hochberg method(Benjamini & Hochberg, 1995). To investigate the robustness of the estimates, mean model coefficients and bias-corrected accelerated percentile bootstrap confidence intervals were calculated from nonparametric bootstrap resampling (1000 iterations) using the ‘boot’ package(Davison & Hinkley, 1997) (version 1.3-25) in R, included in Supplementary Tables. The assumption of linearity was investigated post-hoc using plots of residual values for the top 10 metabolomic and lipidomic differences (ranked by p-value) for each of number of infections, GlycA, and hsCRP.

For secondary analyses to investigate reverse causality, quasi-Poisson regression models(Zeileis, Kleiber, & Jackman, 2008) with each metabolomic/lipidomic measure from cord blood at birth as exposure and total number of infections from birth to 12 months of age as outcome were used, adjusted for infant sex, gestational age, mode of birth, exact age at 12-month time point, birth weight z-score, and maternal household income, and exposure to maternal smoking during pregnancy.

To investigate the potential role of inflammation in mediating associations between infection and metabolomic differences, the ‘mediation’ package(Tingley, Yamamoto, Hirose, Keele, & Imai, 2014) (version 4.5.0) in R was then used to estimate the percentage mediation of 12-month GlycA or hsCRP (as markers of inflammation) for the effect of number of infections on metabolomic and lipidomic differences with an adjusted p-value < 0.1 from the linear regression models described above.

## Results

The flowchart for the 555 infants included in this study is shown in Figure 1, and the cohort characteristics for these infants are shown in Table 1. The median number of total parent-reported infections from birth to 12 months of age was 5 (IQR = [3 to 5]). Median hsCRP and GlycA were 0.25 mg/L [0.08-0.96] and 1.30 mmol/L [1.16-1.48], respectively. Total number of parent-reported infections was more strongly correlated with GlycA (r=0.20) than hsCRP (r=0.11). The distribution of metabolomic and lipidomic measures for the cohort are shown in Supplementary Files 2A and 2B.

**Figure 1.**
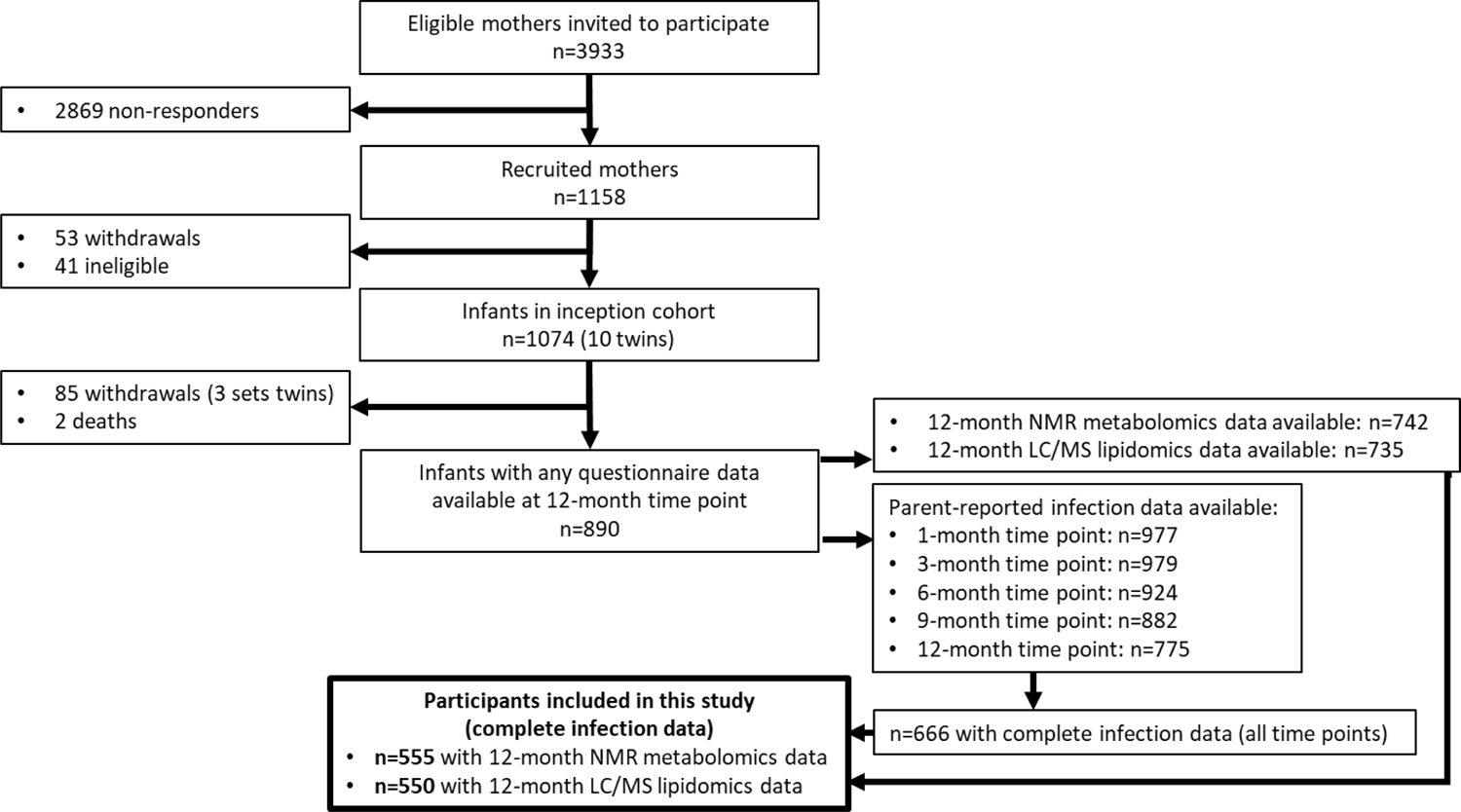
Flowchart of Barwon Infant Study participants included this study (bolded box). Included participants had complete infection data from all 5 time points between birth and 12 months of age, and 12-month plasma NMR metabolomics data. Almost all included participants (n=550 out of 555) had 12-month plasma LC/MS lipidomics data.

**Table 1.** Cohort characteristics (n=555).

| Measure | n (%) |
| --- | --- |
| Sex (female) | 269 (48.4) |
|  | **Mean (SD)** |
| Maternal age at delivery (years) | 31.7 (4.5) |
|  | **n (%)** |
| Maternal smoking during pregnancy (any) | 69 (12.5) |
| Maternal annual household income (AUD) |  |
| <\$25,000 | 11 (2.0) |
| \$25,000 to \$49,999 | 41 (7.5) |
| \$50,000 to \$74,999 | 94 (17.2) |
| \$75,000 to \$99,999 | 145 (26.5) |
| \$100,000 to \$149,999 | 191 (34.9) |
| ≥\$150,000 | 65 (11.9) |
| Mode of birth |  |
| Vaginal | 374 (67.4) |
| Planned caesarean section | 103 (18.6) |
| Unplanned caesarean section | 78 (14.1) |
| Breastfed (any breastfeeding) | 545 (98.2) |
|  | **Median [IQR]** |
| Breastfeeding duration to 52 weeks | 40 [16-52] |
|  | **Mean (SD)** |
| Gestational age (weeks) | 39.5 (1.5) |
| Birth weight (g) | 3538 (521) |
| Age at 12-month time point (months) | 13.0 (0.8) |
| Weight at 12 months of age (kg) | 10.1 (1.3) |
| Weight z-score at 12 months | 0.4 (1.0) |
|  | **Median [IQR]** |
| Number of parent-reported infections | 5 [3-8] |
| GlycA (mmol/L) | 1.30 [1.16-1.48] |
| High-sensitivity C-reactive protein (mg/L) | 0.25 [0.08-0.96] |

### Infection burden and plasma NMR metabolomic profile at 12-months

There was evidence for higher number of infections associating with higher inflammatory markers (GlycA and hsCRP), lower HDL, HDL2, and HDL3 cholesterols, smaller HDL particle size, lower ApoA1, docosahexaenoic acid (DHA), lower total omega-3 fatty acids, lower citrate, higher phenylalanine, and to a lesser extent with higher triglycerides and lower sphingomyelins (Figure 2a, Supplementary File 1C). In models with GlycA as the marker of infection burden, metabolomic differences observed for higher GlycA were largely similar to, but more marked than, those for parent-reported infections; including cholesterols (lower HDL, higher LDL and very-large-density lipoprotein (VLDL) cholesterol), apolipoproteins (lower ApoA1, higher apolipoprotein B (ApoB)), higher total fatty acids, higher total triglycerides and cholines, amino acids (higher phenylalanine, isoleucine, and glycine, lower histidine), glycolysis-related metabolites (higher pyruvate, lower citrate), and lower acetoacetate (Figure 2b, Supplementary File 1D). Bootstrap estimates were generally similar to standard regression estimates for all models.

**Figure 2.**
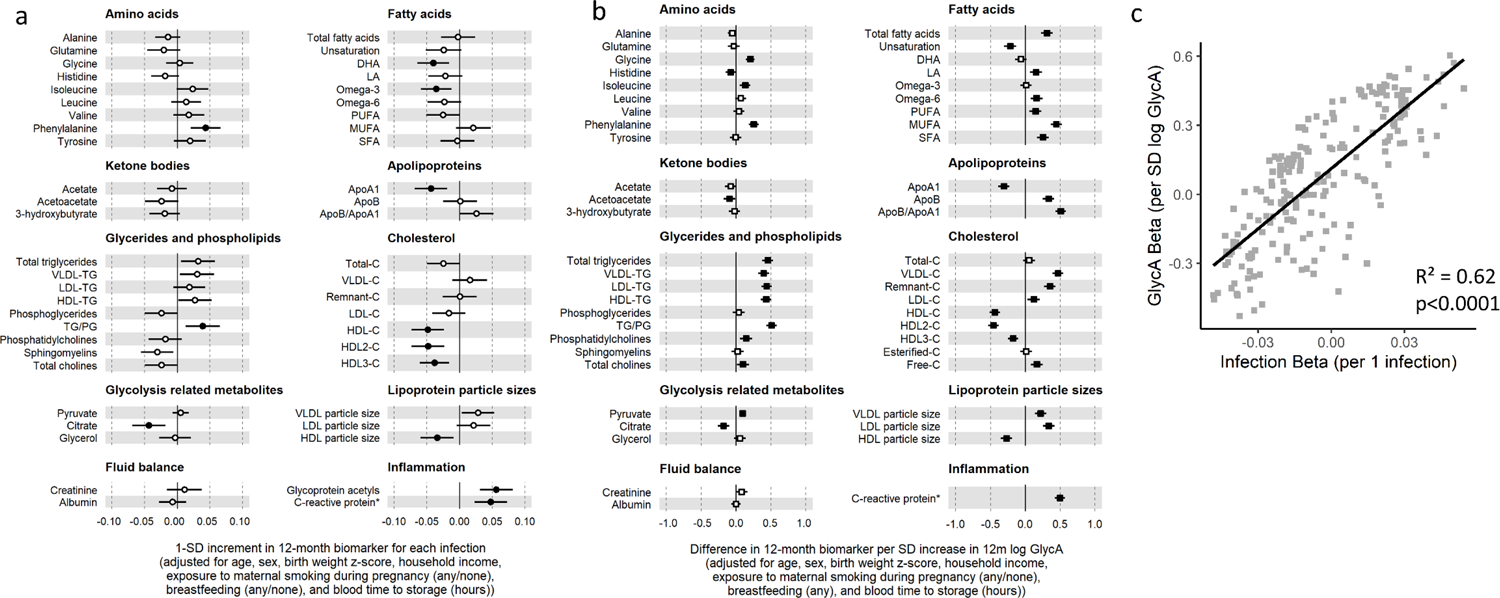
Difference in 12-month plasma NMR metabolomic measures for each increase in parent-reported infection and for each SD increase in GlycA (n=555). Forest plots of the estimated 12-month metabolomic differences for each additional parent-reported infection (a, circle points) or SD log 12-month GlycA (b, square points) from adjusted linear regression models, and the correlation of estimated metabolomic differences for these two exposures (c). Error bars are 95% confidence intervals. Closed points represent adjusted p-value<0.05. Infection exposure model estimates and details for all NMR metabolomic measures are shown in Figure 2 – Source Data 1. GlycA exposure model estimates and details are shown in Figure 2 – Source Data 2.

Higher hsCRP was associated with lower HDL cholesterol, ApoA1 and histidine, and higher phenylalanine, as observed for GlycA, and with lower levels of most other amino acids, total fatty acids, total cholesterols, albumin, phosphoglycerides and VLDL triglycerides, and higher LDL triglycerides (Figure 3a, Supplementary File 1E).

**Figure 3.**
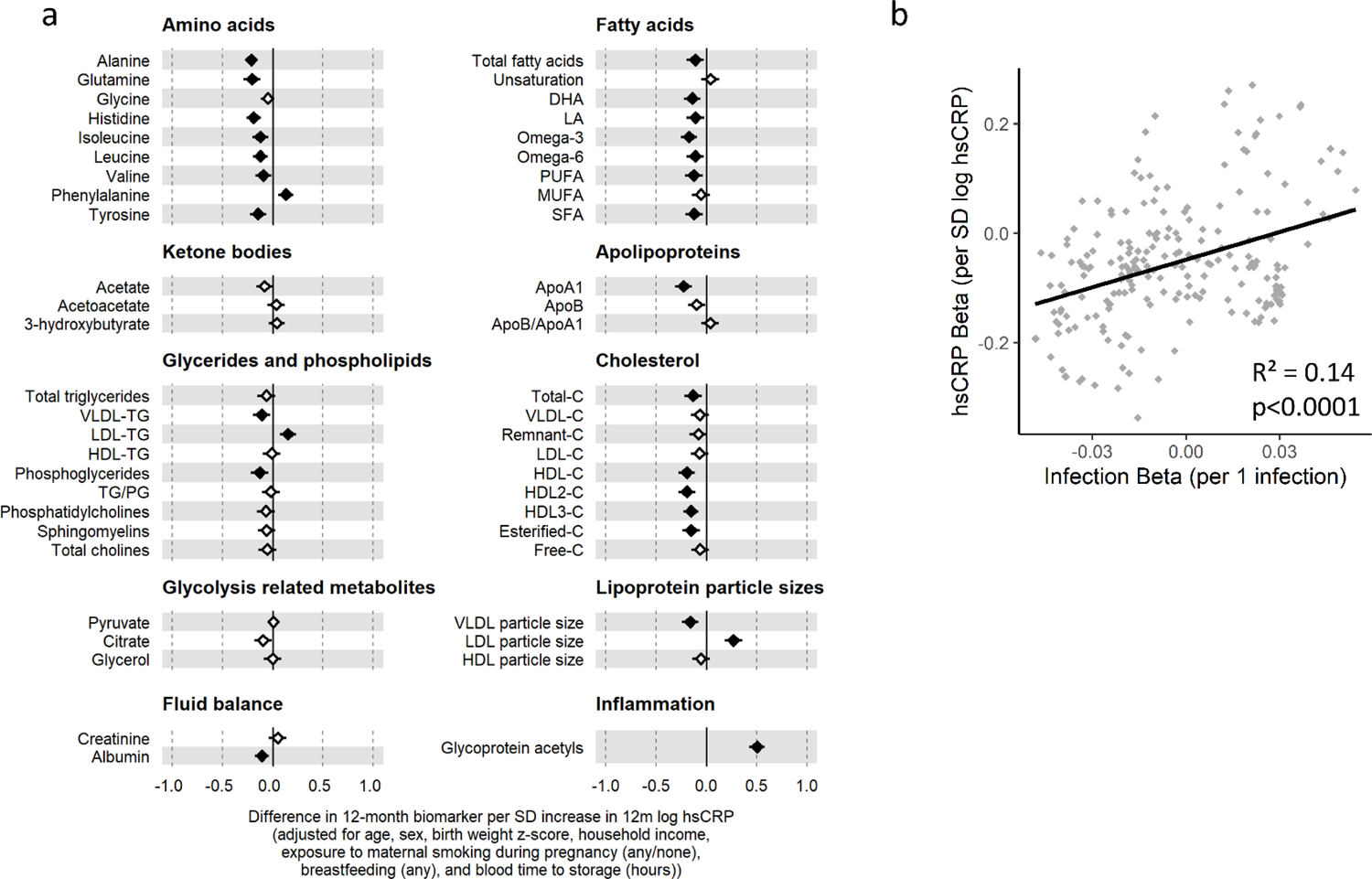
Difference in 12-month plasma NMR metabolomic measures for each SD increase in hsCRP (n=555). Forest plot for the estimated 12-month metabolomic differences for each additional SD log 12-month hsCRP (a, diamond points) from adjusted linear regression models, and the correlation of estimated metabolomic differences for infection and hsCRP (b). Error bars are 95% confidence intervals. Closed points represent adjusted p-value<0.05. hsCRP exposure model estimates and details for all NMR metabolomic measures are shown in Figure32 – Source Data 1.

There was a stronger correlation for the metabolomic differences related to infection and those related to GlycA (r=0.79) than for infection and hsCRP (r=0.37) (Figure 2c, Figure 3b).

In secondary analyses, there was little evidence for associations between serum NMR metabolomic measures at birth at number of parent-reported infections from birth to 12 months of age (Supplementary Files 1F and 3A).

### Infection burden and plasma LC/MS lipidomic profile at 12-months

In regression models with number of parent-reported infections as the marker of infection burden and LC/MS lipids as the outcomes, the 10 lipid measures with the strongest statistical evidence for association with number of infections were hexosylceramides (HexCer(d18:2/18:0), HexCer(d18:2/22:0), HexCer(d16:1/24:0)), trihexosylceramides (Hex3Cer(d18:1/20:0), Hex3Cer(d18:1/22:0), Hex3Cer(d18:1/22:0)), phosphatidylethanolamines (PE(18:0/20:3)), cholesteryl esters (CE(22:5), CE(22:6)) and dehydrocholesterols (DE(18:2)) (Figure 4a, Supplementary File 1G). Considering total lipid concentrations at a class level, infants with more infections had, on average, lower levels of plasmalogens, cholesteryl esters, and dehydrocholesterols.

**Figure 4.**
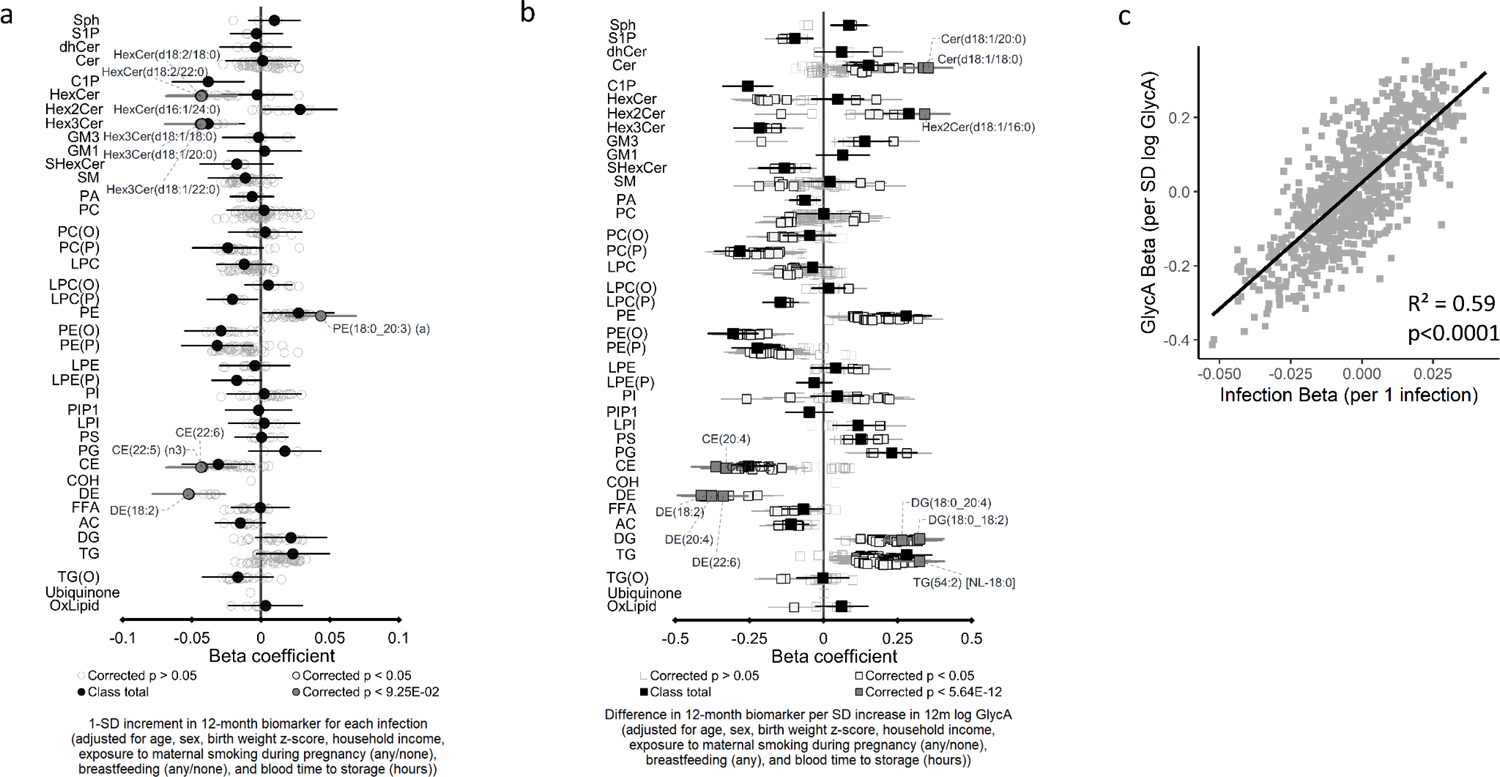
Difference in 12-month plasma LC/MS lipidomic measures for each increase in parent-reported infection and for each SD increase in GlycA (n=550). Forest plots of the estimated 12-month lipidomic differences for each additional parent-reported infection (a, circle points) or SD log 12-month GlycA (b, square points) from adjusted linear regression models, and the correlation of estimated lipidomic differences for these two exposures (c). Error bars are 95% confidence intervals. In (a) and (b), black points represent class totals, closed dark grey points represent the top 10 species differences by p-value, and hollow points with a black outline represent other species with adjusted p<0.05. Infection exposure model estimates and details for all LC/MS lipidomic measures are shown in Figure 4 – Source Data 1. GlycA exposure model estimates and details are shown in Figure 4 – Source Data 2.

Compared to models with number of infections, lipidomic differences were more pronounced when GlycA was considered as the marker of infection burden, including higher sphingomyelins, ceramides, di- and triacylglycerol and phospholipid classes, and lower plasmalogen classes, cholesteryl esters, dehydrocholesterols, and acylcarnitines. (Figure 4b, Supplementary File 1H).

Higher hsCRP was also associated with differences across several lipid classes, particularly lower plasmalogens, lysophosphatidylcholines, and phosphatidylcholine classes (Figure 5a, Supplementary File 1I).

**Figure 5.**
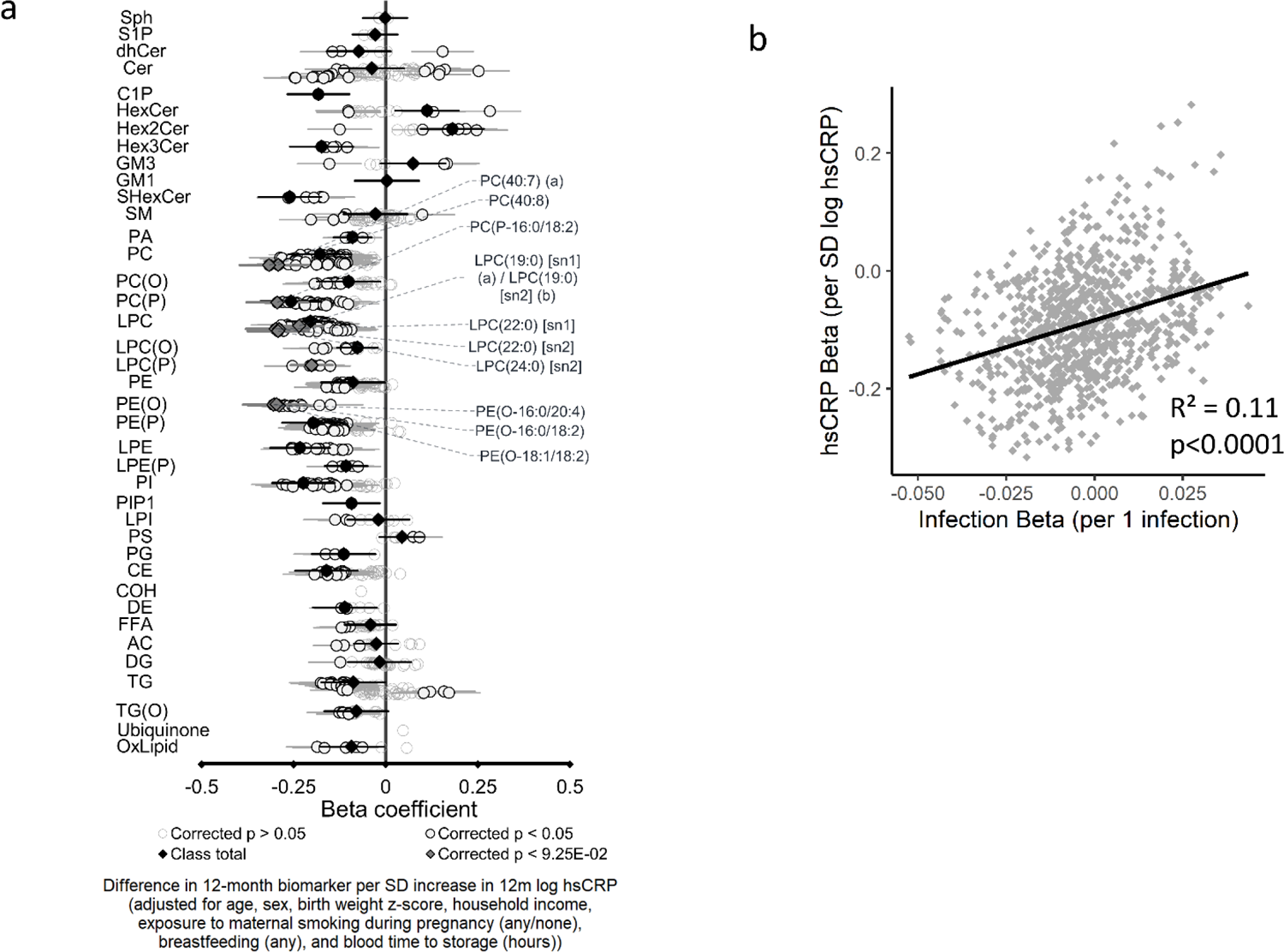
Difference in 12-month plasma LC/MS lipidomic measures for each SD increase in hsCRP (n=550). Forest plot for the estimated 12-month lipidomic differences for each additional SD log 12-month hsCRP (a, diamond points) from adjusted linear regression models, and the correlation of estimated metabolomic differences for infection and hsCRP (b). Error bars are 95% confidence intervals. In (a), black points represent class totals, closed dark grey points represent the top 10 species differences by p-value, and hollow points with a black outline represent other species with adjusted p<0.05. hsCRP exposure model estimates and details for all LC/MS lipidomic measures are shown in Figure 5 – Source Data 1.

There was stronger correlation for lipidomic differences related to infection and GlycA (r=0.77) than for infection and hsCRP (r=0.33) (Figure 4c, Figure 5b).

As with the NMR metabolomic measures, there was little evidence for associations between serum LC/MS lipidomic measures at birth and number of infections from birth to 12 months of age in secondary analyses (Supplementary Files 1J and 3B).

### Mediation analysis

We next assessed whether inflammation (i.e., GlycA or hsCRP at 12 months) mediated the effects of infection on specific metabolite and lipid measures (adjusted p-value < 0.1) (Table 2). For NMR metabolites, the highest estimated mediation by GlycA was for total triglycerides (78.3% (95% CI 40.0%, 334.6%)), triglyceride/phosphoglyceride ratio (70.7% (CI 37.9%, 170.2%)), VLDL triglycerides (70.5% (CI 36.1%, 275.6%)), HDL2-cholesterol (50.6%, (CI 27.9% to 97.8%)), and HDL cholesterol (48.3% (CI 27.3% to 86.9%)). Fatty acids, sphingomyelins and esterified cholesterol showed minimal evidence of mediation by GlycA. In the lipidomic analysis, mediation estimates for GlycA for specific lipidomic measures ranged from 16.9% to 43.5%. There was much lower estimated mediation for inflammation captured by hsCRP; with mediation estimates from 4.7% to 21.6% for metabolomic differences and 4.2% to 21.1% for lipidomic differences.

**Table 2.** Estimated percentage mediation of 12-month GlycA and hsCRP for the effect of number of parent-reported infections on 12-month NMR metabolomic and LC/MS lipidomic differences.

| Measure | Estimated total effect of infections on measure | Adjusted p-value for total effect | Estimated mediation by GlycA* | 95% confidence interval for mediation | P-value for mediation | Estimated mediation by hsCRP* | 95% confidence interval for mediation | P-value for mediation |
| --- | --- | --- | --- | --- | --- | --- | --- | --- |
| **NMR metabolomic** |  |  |  |  |  |  |  |  |
| HDL cholesterol | -0.048 | 0.007 | 48.3% | 27.3% to 86.9% | <0.001 | 16.0% | 5.9% to 36.1% | <0.001 |
| HDL2 cholesterol | -0.048 | 0.007 | 50.6% | 27.9% to 97.8% | <0.001 | 16.0% | 5.5% to 37.6% | 0.002 |
| Phenylalanine | 0.043 | 0.007 | 31.7% | 14.1% to 68.2% | <0.001 | 12.6% | 3.5% to 31.8% | 0.004 |
| Apolipoprotein A1 | -0.043 | 0.012 | 36.6% | 18.3% to 86.7% | <0.001 | 21.6% | 8.8% to 54.8% | <0.001 |
| Citrate | -0.044 | 0.012 | 20.8% | 7.9% to 54.7% | <0.001 | 8.0% | -1% to 28.5% | 0.084 |
| HDL3 cholesterol | -0.038 | 0.013 | 23.3% | 9.1% to 59% | 0.004 | 17.0% | 5.6% to 46% | 0.008 |
| DHA | -0.040 | 0.015 | 5.2% | -6.2% to 20.8% | 0.350 | 14.3% | 2.9% to 39.3% | 0.006 |
| Omega-3 fatty acids | -0.036 | 0.023 | -4.1% | -23.9% to 9.2% | 0.468 | 19.0% | 6.1% to 57.1% | <0.001 |
| Ratio of triglycerides to phosphoglycerides | 0.039 | 0.024 | 70.7% | 37.9% to 170.2% | 0.008 | -4.9% | -26.7% to 7.7% | 0.392 |
| HDL size | -0.034 | 0.049 | 42.9% | 17% to 160.5% | 0.014 | 4.7% | -11.2% to 32.2% | 0.438 |
| Sphingomyelins | -0.031 | 0.071 | -6.5% | -42.9% to 11.1% | 0.352 | 6.3% | -7.5% to 36.1% | 0.276 |
| Total triglycerides | 0.032 | 0.076 | 78.3% | 40% to 334.6% | 0.018 | -11.0% | -62.3% to 3.9% | 0.110 |
| VLDL triglycerides | 0.030 | 0.088 | 70.5% | 36.1% to 275.6% | 0.016 | -18.9% | -100.1% to -3.3% | 0.018 |
| Total esterified cholesterol | -0.029 | 0.088 | -4.4% | -35.1% to 12.2% | 0.518 | 19.1% | 3.2% to 69.9% | 0.024 |
| VLDL size | 0.028 | 0.091 | 40.9% | 14.7% to 168.9% | 0.020 | -29.5% | -187.6% to -7.4% | 0.016 |
| **LC/MS lipidomic** |  |  |  |  |  |  |  |  |
| **Lipid class** |  |  |  |  |  |  |  |  |
| Total DE | -0.053 | 0.003 | 45.1% | 25.8% to 92% | 0.002 | 10.9% | 2.2% to 28.8% | 0.008 |
| Total C1P | -0.038 | 0.059 | 33.3% | 14.6% to 80.5% | 0.002 | 21.1% | 8.1% to 63.7% | 0.002 |
| Total THC | -0.038 | 0.059 | 26.1% | 10.8% to 66.1% | 0.004 | 19.9% | 6.2% to 60.1% | 0.008 |
| **Lipid species** |  |  |  |  |  |  |  |  |
| DE(18:2) | -0.052 | 0.086 | 43.5% | 21.7% to 90.6% | 0.002 | 11.1% | 2.4% to 31.6% | 0.012 |
| HexCer(d16:1/24:0) | -0.042 | 0.092 | 23.3% | 9.5% to 64% | 0.002 | 4.9% | -5.3% to 25.4% | 0.354 |
| HexCer(d18:2/18:0) | -0.044 | 0.092 | 21.9% | 9% to 57.4% | <0.001 | 6.3% | -3.2% to 22.9% | 0.170 |
| HexCer(d18:2/22:0) | -0.043 | 0.092 | 21.7% | 8.1% to 60.4% | <0.001 | 4.3% | -6.3% to 19.5% | 0.396 |
| Hex3Cer(d18:1/18:0) | -0.042 | 0.092 | 21.5% | 8.2% to 49.6% | <0.001 | 15.9% | 4.8% to 39.3% | 0.002 |
| Hex3Cer(d18:1/20:0) | -0.044 | 0.092 | 16.9% | 5.4% to 43.8% | <0.001 | 12.6% | 2.6% to 35.9% | 0.008 |
| Hex3Cer(d18:1/22:0) | -0.043 | 0.092 | 20.7% | 7.5% to 51.8% | <0.001 | 14.9% | 4.5% to 38.5% | 0.004 |
| PE(18:0_20:3) (a) | 0.044 | 0.092 | 31.8% | 14.2% to 78.4% | <0.001 | -7.8% | -29.5% to 1.4% | 0.086 |
| CE(22:5) (n3) | -0.042 | 0.092 | 34.8% | 16.3% to 92.8% | 0.002 | 5.2% | -7.4% to 26.4% | 0.350 |
| CE(22:6) | -0.044 | 0.092 | 32.5% | 16.3% to 74.3% | <0.001 | 7.2% | -2% to 22.2% | 0.140 |
| HexCer(d18:2/24:0) | -0.042 | 0.096 | 19.5% | 5.4% to 62.3% | 0.006 | 5.7% | -5.8% to 23.6% | 0.278 |
| GM3(d18:1/18:0) | -0.043 | 0.096 | 22.0% | 8.6% to 55.1% | <0.001 | 14.6% | 4% to 42% | 0.004 |

\*Mediation percentage is calculated as estimated indirect effect divided by total effect.
Measures shown in Table 2 are the metabolomic and lipidomic measures that were associated with number of parent-reported infection in
linear regression models, as shown in Figure 2 and Figure 4.
Models are adjusted for infant age at sample collection, infant sex, birth weight z-score, maternal household income during pregnancy,
maternal smoking during pregnancy, breastfeeding (any/none), and sample time from collection to storage.

## Discussion

In this study, cumulative parent-reported infection burden from birth to 12 months was associated with adverse NMR metabolomic and LC/MS lipidomic profiles at 12 months of age. Similar but more marked effects on these profiles were evident when considering GlycA, a cumulative inflammation marker, as a surrogate of infection burden. In contrast, differences in metabolomic and lipidomic profiles associated with higher hsCRP were largely distinct from, and less marked than, those for GlycA, suggesting that GlycA may be superior to hsCRP as an early life marker of infection burden. There was evidence that inflammation (GlycA, but not hsCRP) may partly mediate many of the largest metabolomic and lipidomic differences.

The metabolomic profiles observed in this study in infants with greater infection burden (higher triglycerides, lower HDL cholesterol and lower ApoA1) reflect those previously linked to infection, including SARS-CoV-2(Alvarez & Ramos, 1986; Bruzzone et al., 2020; Gallin, Kaye, & O’Leary, 1969; Liuba et al., 2003; Madsen, Varbo, Tybjærg-Hansen, Frikke-Schmidt, & Nordestgaard, 2018), and CVD risk (particularly higher phenylalanine and lower DHA) in adults(Würtz et al., 2015; Würtz et al., 2012). Acute and chronic infection-related metabolic differences have been implicated in atherosclerosis(Khovidhunkit et al., 2000), including increased carotid intima-media thickness(D. P. Burgner, M. A. Sabin, et al., 2015; Liuba et al., 2003) and arterial stiffness(Charakida et al., 2009) in older children, and in adult CVD risk(Bergh et al., 2017; D. P. Burgner, M. N. Cooper, et al., 2015). The mechanisms underpinning the link between inflammation and metabolism are not well characterised, but there is evidence they may share regulatory pathways. For example, lipid-activated nuclear receptors such as peroxisome-proliferator-activated receptors (PPARs) and liver X receptors (LXRs) regulate both lipid metabolism and inflammation(Bensinger & Tontonoz, 2008). Synthesis of leptin, a key regulatory hormone of metabolism, including energy homeostasis(Park & Ahima, 2015), is increased by proinflammatory cytokines during acute infection(Behnes et al., 2012), and is in turn implicated in many inflammatory processes(Abella et al., 2017).

In lipidomic analyses, infection burden was associated with lower levels of several ether phosphatidylethanolamine species (PC(O) and PC(P)), consistent with the reported decrease in these polyunsaturated-fatty acid (PUFA)-containing species in SARS-CoV-2 infected adults(Schwarz et al., 2020). Importantly, these decreases contrast with observed increases in diacyl-phosphatidylethanolamine species (PE), and this discordance in phosphatidylethanolamine species is characteristic of conditions characterised by inflammation, including Alzheimer’s disease(Huynh et al., 2020). The role of phosphatidylethanolamines in infection is incompletely understood, but omega-3 fatty acids (a major class of PUFA) have been implicated in anti-inflammatory pathways(Calder, 2013) and ether phosphatidylethanolamine species are a potential source of PUFAs. Lower trihexosylceramide lipid class, associated with more infections, has been linked to increased BMI and pre-diabetic phenotypes in adults(Meikle et al., 2013; Weir et al., 2013).

The different relationships of the two inflammatory markers GlycA and hsCRP with metabolomic and lipidomic profiles are consistent with a recent study in pregnant women reporting GlycA was more strongly correlated with NMR metabolomic differences than hsCRP(Mokkala et al., 2020). While hsCRP has been widely used as a measure of chronic inflammation, primarily in adults, it is an acute phase reactant that increases rapidly following acute stimulus and returns to baseline levels within a matter of days and is therefore mostly used as a diagnostic adjunct in children with acute infection or inflammation(Gabay & Kushner, 1999). In contrast, there is evidence that GlycA can remain elevated for up to a decade in young adults(Ritchie et al., 2015), and it is considered a superior marker of cumulative inflammation burden, though data of GlycA in early life are sparser. Several studies in adults have reported that these two markers are only moderately correlated(Akinkuolie et al., 2014; Gruppen, Riphagen, et al., 2015), and it is suggested that these markers reflect different (albeit overlapping) inflammatory processes. This is consistent with our findings of distinct metabolomic and lipidomic profiles for these two markers and reflects other findings that show different relationships of GlycA and hsCRP with cardiovascular and metabolic phenotypes.

For example, GlycA is independently associated with risk of CVD and with enzymatic esterification of free cholesterol, even after adjustment for hsCRP(Duprez et al., 2016; Gruppen, Connelly, Otvos, Bakker, & Dullaart, 2015; Muhlestein et al., 2018). Associations between GlycA and lipolysis rates(Levine et al., 2020) and gut microbiome diversity(Mokkala et al., 2020) are also stronger than those reported for hsCRP.

### Strengths and limitations

This large prospective cohort study is the first to examine the relationship between infection, inflammation and plasma NMR metabolomic and LC/MS lipidomic profiles in early life, with implications for later CVD risk. Findings were consistent using either parental-reported infections (potentially prone to reporting bias) or 12-month GlycA as a measure of cumulative inflammatory burden. We found no evidence that birth plasma metabolomic profiles were associated with infection burden in the first 12 months of life (i.e., reverse causation; Supplementary Files 3A and 3B).

Limitations include a single measure of the metabolome and lipidome as an outcome at 12 months of age. Although mediation analyses used cross-sectional data, GlycA is a marker of cumulative inflammation(Collier et al., 2019; Ritchie et al., 2015) and reflects inflammatory events occurring prior to the 12-month time point. Despite rigorous adjustment, there may be unmeasured confounding. We considered total parent-reported infections, as defining clinical categories of infection given the non-specific symptoms and signs in infancy is challenging. Validation of all parent-reported infections was not feasible and only a minority (70 infections in 555 infants) resulted in medical attention(Rowland et al., 2020). However, the strong correlation between metabolomic and lipidomic differences for number of reported infections and for GlycA suggest that parent report is a reasonable measure of infection-induced inflammatory burden. Most childhood infections are viral, and microbial testing is impractical in non-hospitalised children and cannot necessarily differentiate between colonisation and infection. Notwithstanding, distinct lipid differences have been reported for children with bacterial versus viral infections(X. Wang et al., 2019), and different infectious aetiologies in adults have been linked to differential CVD risk(Cowan et al., 2020). Finally, the relative lack of racial/ethnic diversity in our cohort may limit generalisability of our findings.

## Conclusions

In summary, we present evidence for higher infection burden in early life leading to pro-atherogenic and pro-diabetic plasma metabolome and lipidome at 12 months of age, and for inflammation partly mediating these relationships. GlycA may be a better marker of early life infection burden than hsCRP. These findings suggest that the impact of the cumulative infection and inflammation burdens previously implicated in adult cardiometabolic disease may begin in infancy, thereby offering opportunities for early prevention. Further work is required to determine the potential consequences these adverse metabolomic profiles in early life have on later risk of disease, and how the relationships between infections, inflammation and metabolomic and lipidomic profiles might differ across age groups, pathogen, and clinical severity of infection.

## Data Availability

The data used in this study are available upon reasonable request with approval from BIS data custodians. Interested parties can contact the corresponding author.

## Acknowledgements

The authors thank the BIS participants for the generous contribution they have made to this project. The authors also thank current and past staff for their efforts in recruiting and maintaining the cohort and in obtaining and processing the data and biospecimens. The members of the BIS Investigator Group are the following: Peter Vuillermin, Anne-Louise Ponsonby, John Carlin, Mimi LK Tang, Amy Loughman, Toby Mansell, Lawrence Gray, Martin O’Hely, Richard Saffery, Sarath Ranganathan, David Burgner and Peter Sly. We thank Terry Dwyer and Katie Allen for their past work as foundation investigators.

## Data availability

With the approved ethics for this study, the individual participant data cannot be made freely available online. Interested parties can access the data used in this study upon reasonable request, with approval by the Barwon Infant Study data custodians. As part of this process, researchers will be required to submit a project concept for approval, to ensure the data is being used responsibly, ethically, and for scientifically sound projects.

Source data files have been uploaded for each of the results figures (Figures 2 to 5) showing the model summary data for all metabolomic and lipidomic measures, including those not presented in figures.

Source code for all analyses have been uploaded as Source Code 1 and 2.

## Source and Supplementary Data

**Figure 2 – Source Data 1.** Summary of regression models for difference in NMR metabolomic measures per 1 increase in parent-reported infection, adjusted for any breastfeeding.

**Figure 2 – Source Data 2.** Summary of regression models for difference in NMR metabolomic measures per SD increase in log 12-month GlycA, adjusted for any breastfeeding.

**Figure 3 – Source Data 1.** Summary of regression models for difference in NMR metabolomic measures per SD increase in log 12-month hsCRP, adjusted for any breastfeeding.

**Figure 4 – Source Data 1.** Summary of regression models for difference in LC/MS lipidomic measures per 1 increase in parent-reported infection, adjusted for any breastfeeding.

**Figure 4 – Source Data 2.** Summary of regression models for difference in LC/MS lipidomic measures per SD increase in log 12-month GlycA, adjusted for any breastfeeding.

**Figure 5 – Source Data 1.** Summary of regression models for difference in LC/MS lipidomic measures per SD increase in log 12-month hsCRP, adjusted for any breastfeeding.

**Supplementary File 1**. Secondary and sensitivity analyses.

A. Summary of regression models for difference in NMR metabolomic measures for infections, GlycA and hsCRP for participants with plasma processing time less than 4 hours.
B. Summary of regression models for difference in LC/MS lipidomic measures for infections, GlycA and hsCRP for participants with plasma processing time less than 4 hours.
C. Summary of regression models for difference in NMR metabolomic measures per 1 increase in parent-reported infection, adjusted for duration of breastfeeding.
D. Summary of regression models for difference in NMR metabolomic measures per SD increase in log 12-month GlycA, adjusted for duration of breastfeeding.
E. Summary of regression models for difference in NMR metabolomic measures per SD increase in log 12-month hsCRP, adjusted for duration of breastfeeding.
F. Summary of regression models for difference in log number of parent-reported infections per SD increase in log birth NMR metabolomic measure.
G. Summary of regression models for difference in LC/MS lipidomic measures per 1 increase in parent-reported infection, adjusted for duration of breastfeeding.
H. Summary of regression models for difference in LC/MS lipidomic measures per SD increase in log 12-month GlycA, adjusted for duration of breastfeeding.
I. Summary of regression models for difference in LC/MS lipidomic measures per SD increase in log 12-month hsCRP, adjusted for duration of breastfeeding.
J. Summary of regression models for difference in log number of parent-reported infections per SD increase in log birth LC/MS metabolomic measure.

**Supplementary File 2**. Cohort distribution of NMR metabolomic and LC/MS lipidomic measures.

A. Distribution of NMR metabolomic measures in cohort.
B. Distribution of LC/MS lipidomic measures in cohort.

**Supplementary File 3**. Forest plots of models investigating reverse causation: models with NMR metabolomic and LC/MS lipidomic measures at birth as the exposures and number of parent-reported infections as the outcome.

A. Difference in log number of parent-reported infections from birth to 12 months of age for each SD increase in log serum NMR metabolomic measure at birth.
B. Difference in log number of parent-reported infections from birth to 12 months of age for each SD increase in log serum LC/MS lipidomic measure at birth.

**Source Code 1**. Code (R Script) for all analyses of NMR metabolomics data in this study.

**Source Code 2**. Code (R Script) for all analyses of LC/MS lipidomics data in this study.

